# Intermittent calorie restriction alters T cell subsets and metabolic markers in people with multiple sclerosis

**DOI:** 10.1101/2022.01.11.22269094

**Authors:** Kathryn C. Fitzgerald, Pavan Bhargava, Matthew D. Smith, Diane Vizthum, Bobbie Henry-Barron, Michael D. Kornberg, Sandra D. Cassard, Dimitrios Kapogiannis, Patrick Sullivan, David J. Baer, Peter A. Calabresi, Ellen M. Mowry

## Abstract

**Background:** Intermittent fasting or calorie restriction (CR) diets provide anti-inflammatory and neuroprotective advantages in models of multiple sclerosis (MS); data in humans are sparse.

**Methods:** We conducted a randomized-controlled feeding study of different CR diets in 36 people with MS over 8 weeks. Patients were randomized to receive either: a daily CR diet (22% reduction in calories, 7 days/week), an intermittent CR diet (75% reduction, 2 days/week; 100%, 5 days/week), or a weight-stable diet (100%, 7 days/week). Untargeted metabolomics was performed on plasma samples at weeks 0, 4 and 8 at Metabolon Inc (Durham, NC). Flow cytometry of cryopreserved peripheral blood mononuclear cells at weeks 0 and 8 were used to identify CD4^+^ and CD8^+^ T cell subsets including effector memory, central memory, and naïve cells.

**Results:** 31 (86%) completed the trial. Over time, individuals randomized to intermittent CR had significant reductions in CD4^+^_CM_ -4.87%; 95%CI: -8.59%, -1.15%; p=0.01), CD4^+^_EM_ (−3.82%; 95%CI: -7.44, - 0.21; p=0.04), and CD8^+^_EM_ (−6.96%; 95%CI: -11.96, -1.97; p=0.006) with proportional increases in naïve subsets (CD4^+^_Naïve_: 5.81%; 95%CI: -0.01, 11.63%; p=0.05; CD8^+^_Naïve_: 10.11%; 95%CI: 3.30, 16.92%; p=0.006). No changes were observed for daily CR or weight-stable diets. Larger within-person changes in lysophospholipid and lysoplasmalogen metabolites in intermittent CR were associated with larger reductions in memory T cell subsets and larger increases in naïve T cell subsets.

**Conclusions:** In people with MS, an intermittent CR diet was associated with reduction in memory T cell subsets. The observed changes may be mediated by changes in specific classes of lipid metabolites.

**Trial Registration:** This study is registered on Clinicaltrials.gov with identifier NCT02647502.

**Funding:** National MS Society, NIH, Johns Hopkins Catalyst Award

## Introduction

Multiple sclerosis (MS) is an autoimmune and neurodegenerative disorder of the central nervous system (CNS).(1) Progression of disability is currently irreversible, and prognosis is highly variable, highlighting the need for continued research regarding modifiable risk and protective factors for disease worsening. The role of diet has emerged as an important factor that may modulate MS course.(2–5) Diet could potentially do so through direct modulation of the immune system, via alterations to gut bacteria, or by changes to metabolism (e.g., modification of oxidative stress or mitochondrial function).

One aspect of diet that shows promise as an interventional target involves the timing and amount of caloric intake. Intermittent calorie restriction (CR) or fasting prior to the induction of experimental autoimmune encephalomyelitis (EAE), a mouse model of MS, leads to a less aggressive course.(6–8) Intermittent CR also reduces pro-inflammatory cytokines and other inflammatory markers in EAE mice, and emerging studies suggest intermittent CR promotes regeneration of oligodendrocytes in other mouse models of MS.(6–8) In humans who are healthy, obese, or have asthma, intermittent CR is associated with similar reductions in inflammatory markers and may have stronger metabolic effects than spreading the same calorie deficit over the course of a week.(9–11)

We compared different types of CR diets with a weight-stable diet in a controlled feeding study of people with MS entitled the “Alternating the Timing and Amount of Calories in MS” or ATAC-MS. CR diets included an intermittent CR or a ‘fasting-style’ diet with a more traditional daily CR in which calorie deficits are distributed evenly over a 7-day period. We found that both types of CR diets are not only safe and feasible but are also associated with weight loss and improvements in emotional health.(3) Herein, we extend these initial results by evaluating how different types of CR diets impact more proximal biologic mediators with specific relevance to MS, including changes in adipokines (e.g., leptin, adiponectin)(12–14), immune cell subsets,(15–17) and the plasma metabolome.(18, 19) We also explore whether within-person changes in metabolite levels potentially mediate changes in metabolic traits or immune cell subsets. Taken together, these results may further provide a valuable step forward in understanding the effects of different types of CR on important intermediate MS outcomes.

## Results

### Characteristics of Study Participants

People with MS were randomized to receive 1 of 3 diets: a daily CR diet (22% daily reduction in energy needs), an intermittent CR diet (75% reduction in calorie needs 2 days/week; 100% of daily needs 5 days/week), and a weight-stable diet (100% of daily calorie needs); an overview of the study design is provided in **Figure 1a**. Baseline characteristics of the included 36 participants are provided in **Table 1** and were generally comparable across the three study diets. Participants on average were aged 37.4 (standard deviation [SD]: 7.4), were predominantly female (81%), and had an average body mass index (BMI) of 32.6 kg/m^2^. Of the 36 participants enrolled, 31 (86%) completed the 8-week feeding portion of the study. One participant withdrew from the intermittent CR arm, one participant withdrew from the daily CR, and three participants withdrew from the control arm (**Figure 1b**). As previously reported, participants randomized to the intermittent CR lost on average 3.0kg (interquartile range [IQR]: -2.0kg, - 4.1kg), and participants randomized to daily CR lost on average 3.6kg (IQR: -3.0kg, -4.1kg); weight loss did not differ significantly between the two types of CR (3).

**Figure 1.**
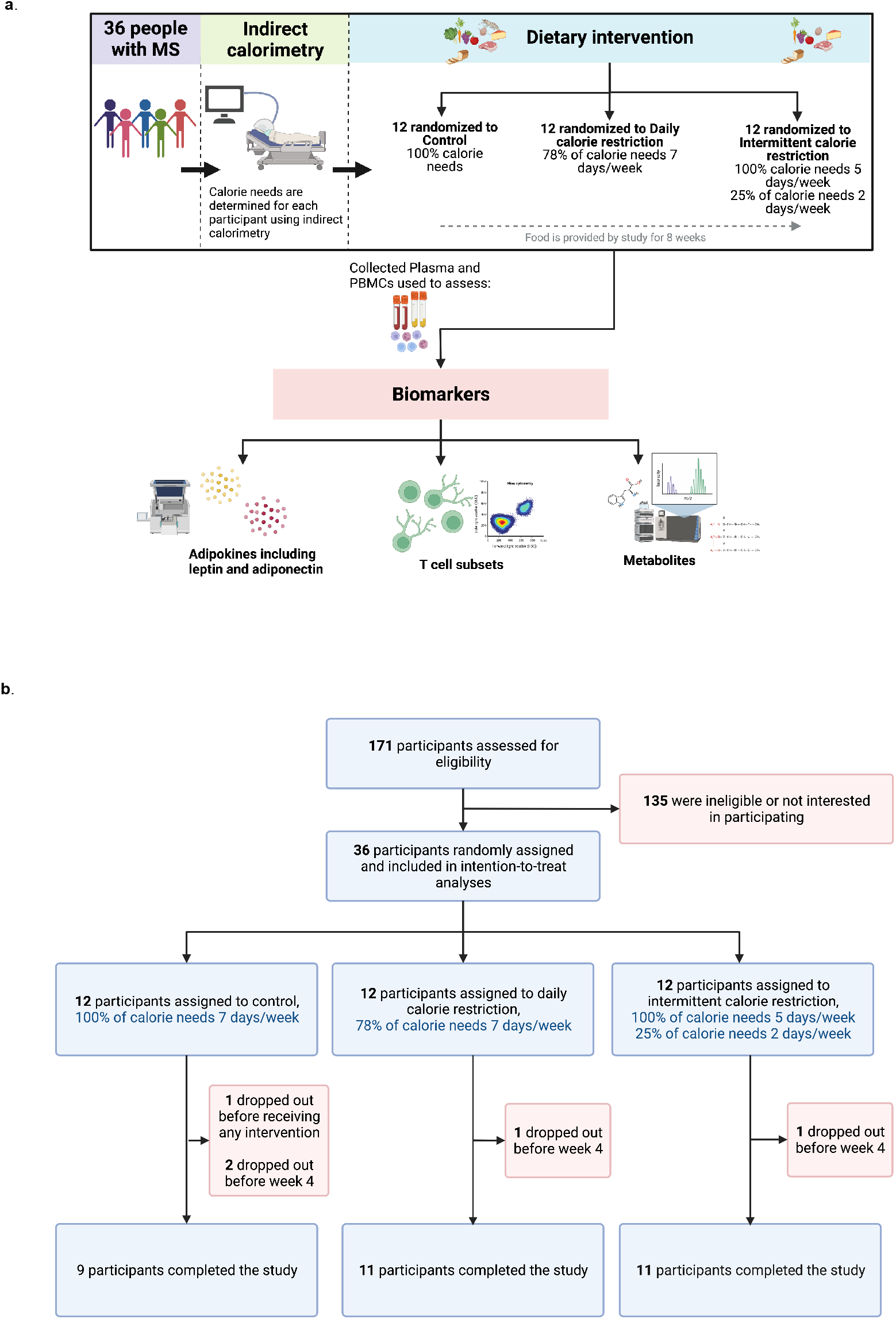
**a**. Overview of the ATAC-MS study design. **b**. Consort diagram for ATAC-MS participants. Figure 1 (both **a** and **b**) was Created with BioRender.com

**Table 1.** Demographic and Clinical Characteristics of ATAC-MS Study Participants.

|  | Intermittent | Daily | Control |
| --- | --- | --- | --- |
| N | 12 | 12 | 12 |
| Age, years, mean (SD) | 38.50 (7.38) | 40.50 (5.44) | 33.33 (6.98) |
| Males, n (%) | 2 (16.7) | 2 (16.7) | 3 (25.0) |
| Black race, n (%) | 3 (25.0) | 4 (33.3) | 3 (25.0) |
| Hispanic/Latino, n (%) | 1 (8.3) | 0 (0.0) | 2 (16.7) |
| EDSS, mean (SD) | 1.75 (0.72) | 1.67 (0.91) | 1.08 (1.14) |
| Disease modifying therapy |  |  |  |
| glatiramer acetate | 7 (58.3) | 8 (66.7) | 4 (33.3) |
| interferon beta | 4 (33.3) | 3 (25.0) | 6 (50.0) |
| none | 1 (8.3) | 1 (8.3) | 2 (16.7) |
| Adherence*, %, mean (SD) | 23.09 (36.11) | -18.87 (28.06) | -11.44 (19.35) |
| BMI, kg/m<sup>2</sup>, mean (SD) |  |  |  |
| Baseline | 31.62 (6.96) | 35.13 (10.13) | 32.41 (7.20) |
| End of study | 30.70 (6.84) | 33.64 (9.86) | 31.99 (7.68) |
| Adiponectin, µg/mL, mean (SD) |  |  |  |
| Baseline | 19042.26 (11579.11) | 17320.07 (15839.92) | 18808.08 (6579.05) |
| End of study | 18942.17 (11300.58) | 15864.67 (13579.47) | 15278.04 (8605.42) |
| Leptin, µg/mL, mean (SD) |  |  |  |
| Baseline | 16310.42 (20259.71) | 27260.94 (24928.00) | 31015.69 (28682.91) |
| End of study | 26894.88 (39094.44) | 27920.59 (23921.98) | 19997.37 (19699.90) |

\*Calculated as the difference between percentage calories consumed versus provided by study at week 8.

### No Change in Adipokine Levels with CR

We did not observe changes in serum leptin or adiponectin in either of the CR diets over the 8-week period. Unexpectedly, both leptin and adiponectin decreased in the control arm (**Figure 2a**; **Table 2;** difference in geometric mean over time: leptin: -0.93µg/mL; 95% CI: -1.87 to 0.00; p=0.05; adiponectin: - 0.54µg/mL; 95% CI: -0.86, -0.21; p=0.001).

**Figure 2.**
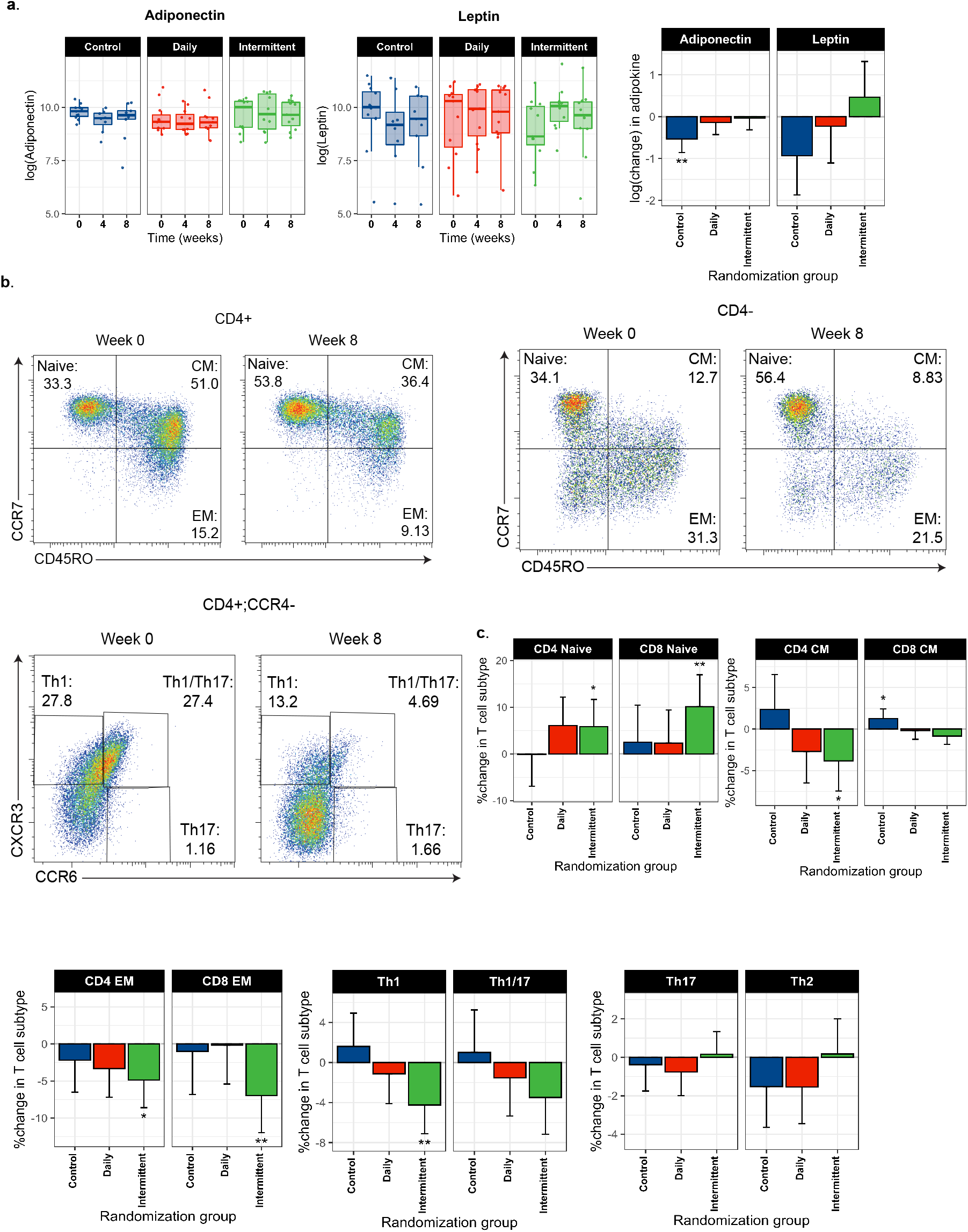
**a**. Boxplots depicting log-transformed leptin and adiponectin values over the course of the study for each of the study diets. The final bar chart depicts the rate of change in adipokines over the course of the study for each of the study diets. **b**. Representative flow cytometry plots for selected T cell subsets for week 0 and week 8 for individuals randomized to intermittent CR. **c**. Rate of change in T cell subsets over the 8-week period across the three diets. Rates were additionally adjusted for age, sex, disease modifying therapy and adherence to provided diets (calculated as the difference in calories consumed versus calories provided by the study). For all bar plots, * denotes unadjusted p<0.05 and ** denotes unadjusted p<0.01.

**Table 2.** Change in metabolic traits and immune cell subsets.

|  | Change over 8-week period |  |  |  |  |  | Daily vs.<br>Control | Intermittent<br>vs. Control | Intermittent<br>vs. Daily |
| --- | --- | --- | --- | --- | --- | --- | --- | --- | --- |
|  | Intermittent |  | Daily |  | Control |  |  |  |  |
|  | log(Change) (95% CI) | P value | log(Change) (95% CI) | P value | log(Change) (95% CI) | P value | P value | P value | P value |
| **Metabolic trait\*** |  |  |  |  |  |  |  |  |  |
| Leptin, µg/mL | 0.46 (-0.39, 1.32) | 0.29 | -0.23 (-1.11, 0.65) | 0.61 | -0.93 (-1.87, 0.00) | 0.05 | 0.04 | 0.28 | 0.26 |
| Adiponectin, µg/mL | -0.03 (-0.31, 0.25) | 0.82 | -0.14 (-0.43, 0.15) | 0.34 | -0.54 (-0.86, -0.21) | 0.001 | 0.03 | 0.61 | 0.06 |
| **Immune Cell\*** |  |  |  |  |  |  |  |  |  |
|  | % Change (95% CI) | P value | % Change (95% CI) | P value | % Change (95% CI) | P value | P value | P value | P value |
| CD4 CM | -3.82% (-7.44%, -0.21%) | 0.04 | -2.70% (-6.48%, 1.09%) | 0.16 | 2.35% (-1.87%, 6.56%) | 0.28 | 0.04 | 0.68 | 0.07 |
| CD4 EM | -4.87% (-8.59%, -1.15%)** | 0.01 | -3.33% (-7.21%, 0.56%) | 0.09 | -2.18% (-6.51%, 2.14%) | 0.32 | 0.38 | 0.58 | 0.68 |
| CD4 Naive | 5.81% (-0.01%, 11.63%) | 0.05 | 6.06% (-0.03%, 12.15%) | 0.05 | -0.15% (-6.93%, 6.64%) | 0.97 | 0.21 | 0.96 | 0.16 |
| CD8 CM | -0.85% (-1.86%, 0.16%) | 0.10 | -0.18% (-1.24%, 0.87%) | 0.73 | 1.26% (0.09%, 2.44%) | 0.03 | 0.02 | 0.39 | 0.06 |
| CD8 EM | -6.96% (-11.96%, -1.97%)** | 0.006 | -0.18% (-5.40%, 5.04%) | 0.95 | -1.00% (-6.82%, 4.82%) | 0.74 | 0.15 | 0.07 | 0.83 |
| CD8 Naive | 10.11% (3.30%, 16.92%)** | 0.004 | 2.30% (-4.80%, 9.41%) | 0.52 | 2.51% (-5.40%, 10.42%) | 0.53 | 0.18 | 0.13 | 0.97 |
| Th1 | -4.26% (-7.11%, -1.40%)** | 0.004 | -1.12% (-4.10%, 1.85%) | 0.46 | 1.61% (-1.71%, 4.93%) | 0.34 | 0.02 | 0.15 | 0.21 |
| Th1/17 | -3.49% (-7.16%, 0.18%) | 0.06 | -1.53% (-5.35%, 2.28%) | 0.43 | 1.00% (-3.25%, 5.25%) | 0.65 | 0.14 | 0.48 | 0.36 |
| Th2 | 0.18% (-1.65%, 2.00%) | 0.85 | -1.55% (-3.45%, 0.35%) | 0.11 | -1.52% (-3.64%, 0.59%) | 0.16 | 0.25 | 0.21 | 0.99 |
| Th17 | 0.15% (-1.03%, 1.33%) | 0.8 | -0.76% (-1.99%, 0.47%) | 0.23 | -0.38% (-1.75%, 0.99%) | 0.59 | 0.58 | 0.31 | 0.67 |

\*Denotes the rate of change over 8 weeks in metabolic trait or T cell subset. Differences are additionally adjusted for age, sex, MS disease modifying therapy, and adherence to study diets.
\*\*P-values for immune cell analyses were adjusted for false discovery; \*\* denotes FDR-adjusted p-values <0.05

### Intermittent CR alters T cell subsets

Over time, individuals randomized to intermittent CR had significant reductions in memory T cell subsets including CD4^+^_EM_ (**Figure 2b** and **2c**; **Table 2**; -4.87%; 95% CI: -8.59%, -1.15%; FDR-adjusted p=0.01) and CD8 ^+^_EM_ (−6.96%; 95% CI: -11.96, -1.97; FDR-adjusted p=0.03) with concomitant increases in naïve subsets (CD8^+^_Naïve_: 10.11%; 95% CI: 3.30, 16.92%; FDR-adjusted p=0.02). Individuals randomized to intermittent calorie restriction also experienced significant reductions in Th1 cells (−4.26%; 95% CI: -7.11%, -1.40%; FDR-adjusted p=0.02) and trends toward a potential reduction in CD4^+^_CM_ (−3.82%; 95% CI: -7.44, -0.21; FDR-adjusted p=0.09) and Th1/17 cells (−3.49%; 95% CI: -7.16%, 0.18%; FDR-adjusted=0.11) and increase in (CD4^+^_Naïve_: 5.81%; 95% CI: -0.01%, 11.63%; FDR-adjusted p=0.10) over the 8-week follow-up. No notable changes in T cell subsets were observed for individuals randomized to daily CR or weight stable diets.

### CR alters the circulating metabolome

In metabolomics analyses, we noted changes in several different classes of lipid metabolites. For example, in pathway-based analyses using metabolite-set enrichment analysis (MSEA; an adaptation of gene-set enrichment analysis for metabolomics data; see **Methods**), we noted general increases in acyl carnitine metabolites (which shuttle fatty acids into mitochondria) for both CR diets. The normalized expression scores (NES), which are composite measures incorporating individual statistical tests for each of the metabolites considered and normalized by the size of the pathway, were >2-fold higher for each diet (for any CR: NES=2.61, FDR-adjusted p=8.37E-09; for intermittent CR: NES=2.73, FDR-adjusted p=3.20E-06; for daily CR: NES=2.49, FDR-adjusted p=1.25E-06; **Figure 3**). We also noted general reductions in glycerophospholipids in individuals randomized to CR, with notable changes in phosphatidylcholines (major constituents of plasma membranes), plasmalogens (endogenous antioxidants) and lysoplasmalogens (products of plasmalogen metabolism). These differences were particularly notable in individuals randomized to intermittent CR, in which significant changes were observed predominantly for phosphatidylethanolamine (PE) plasmalogens rather than phosphatidylcholine (PC) plasmalogens (**Figure 3c**). CR diets were also associated with increases in glycine, threonine, and serine metabolism (NES: 1.91; FDR-adjusted p=0.05). In sensitivity analyses, we observed relatively consistent results when we applied our rank-based pathway analysis. Results for all metabolites over time and metabolic pathways are summarized in **Supplemental Table 1**. Results for individual metabolites organized by pathways are also provided as a part of the accompanying browser available at https://brbdai-kathryn-fitzgerald.shinyapps.io/Calorie-restriction-biomarker/.

**Figure 3.**
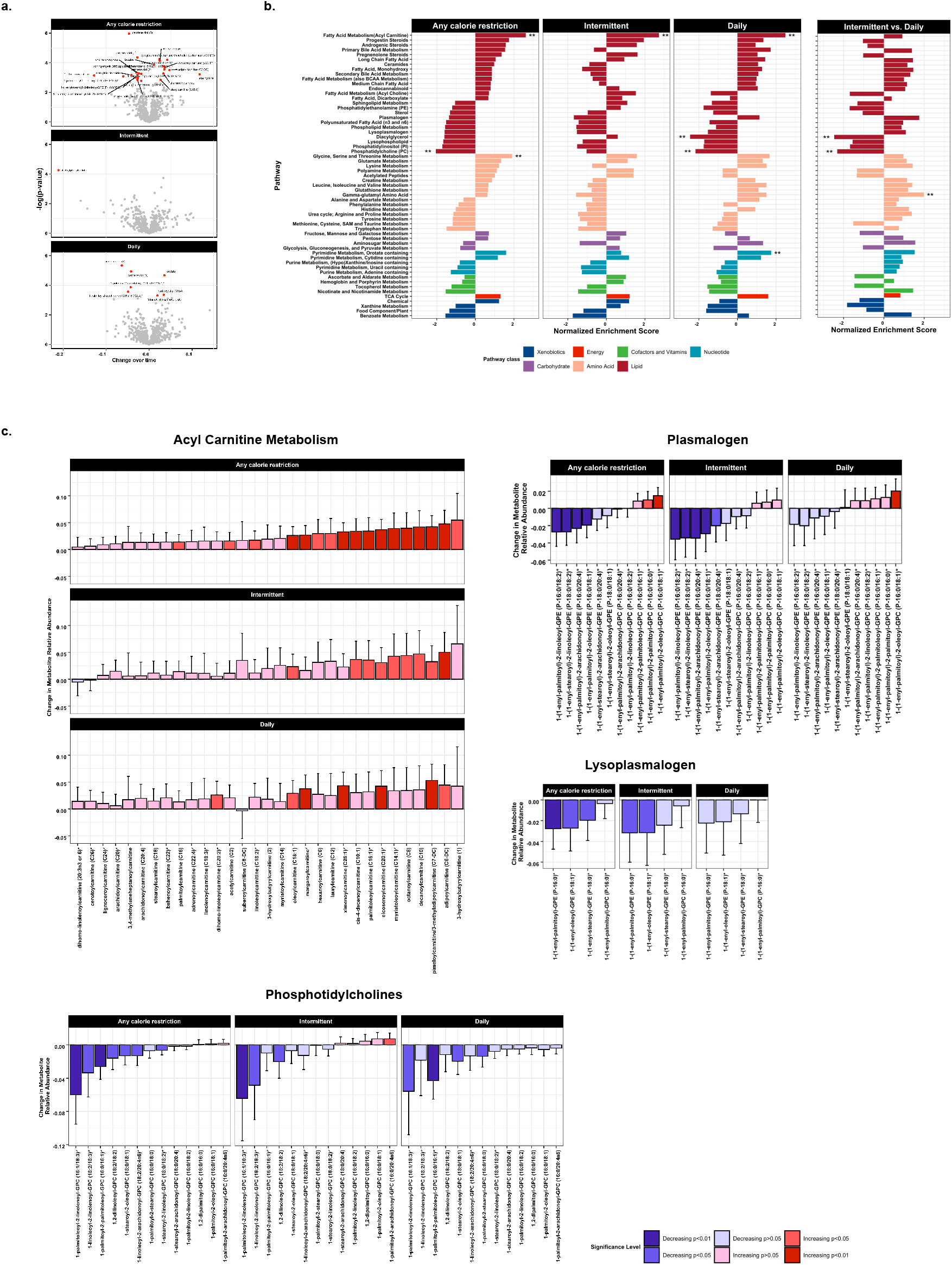
**a**. Volcano plot depicting results of the individual metabolite analyses. Red labeled metabolites denote metabolites that changed significant (FDR-adjusted p-value <0.05). Rates were additionally adjusted for age, sex, disease modifying therapy and adherence to provided diets (calculated as the difference in calories consumed versus calories provided by the study. **b**. Normalized expression scores (NES) from metabolite set enrichment analyses (MSEA) derived for metabolite changes over time for any CR, intermittent and daily CR. The last panel denotes the differences in NES across pathways between intermittent versus daily CR. ** denotes pathways which were significant after accounting for false discovery (FDR-adjusted p-value <0.05). **c**. Individual results for metabolites in selected pathways identified in MSEA. Bars are colored based on their direction and level of significance in individual analyses.

The rate of change for each metabolite for individuals randomized to intermittent CR was positively correlated with the rate of change for each metabolite for individuals randomized to daily CR, suggesting generally similar change in the metabolome for each of the CR diets. For example, the Spearman correlation between pathway NES for intermittent CR versus NES for daily CR was 0.86 (95% CI: 0.76, 0.92; p <1E-16; **Figure 3b**), and the Spearman correlation between rates of change between individual metabolites for individuals randomized to intermittent CR and individuals randomized to daily CR was 0.30 (95% CI: 0.22, 0.39).

### Within-Person Change in Metabolites due to CR predict change in Immune Cell Subsets

In exploratory analyses, we also assessed whether changes in adipokines or immune cell subtypes were related to within-person changes occurring in the plasma metabolome. We concentrated these analyses on individuals randomized to the intermittent CR as these individuals, on average, exhibited significant changes in immune cell subsets (relative to those randomized to daily or control diets). Interestingly, in individuals randomized to intermittent CR, we noted changes in products of glycerophospholipid metabolism that may mediate some of the observed changes in immune cell subsets (**Figure 4a, 4b** and **4c, Supplemental Table 2** and our browser: <https://brbdai-kathryn-fitzgerald.shinyapps.io/Calorie-restriction-biomarker/>). For example, within-person reductions in lysophospholipids were generally associated with concomitant reductions in in CD4+_CM_ (**Figure 4a**; NES: -1.93; FDR-adjusted p=0.005), CD8+_CM_ (NES: -2.16; FDR-adjusted p=0.001), Th1 (NES: -1.87; FDR-adjusted p=0.003) and Th1/17 (NES: - 2.26; FDR-adjusted p=8.89E-5) subsets and increase in the CD4+_Naïve-_ subset (NES: 2.07; FDR-adjusted p=0.006). Within-person reductions in lysophospholipids were also associated with increases in CD4+_Naïve-_ subsets (NES: 2.07; FDR-adjusted p=0.006). Within-person change in lysolipids was also associated with increases in adiponectin (NES: 2.39; FDR-adjusted p=1.01E-4) and decreases in leptin (NES: 3.14; FDR-adjusted p=6.86E-7) levels. Similarly, within-person reductions in lysoplasmalogens were associated with reductions in CD4+_EM_ (NES: -1.57; FDR-adjusted p=0.04), CD8+_EM_ (NES: -1.57; FDR-adjusted p=0.04), Th1 (NES: -1.93; FDR-adjusted p=0.005), and Th1/17 (NES: -1.70; FDR-adjusted p=0.01). Notably, similar within-person changes in lysophospholipids or lysoplasmalogens were not associated with changes in T cell subsets for individuals randomized to daily CR (**Supplemental Figure 1**). Within-person change in leptin or adiponectin levels were not associated with changes in T cell subsets (all FDR-adjusted p values >0.05; **Supplemental Table 3**).

**Figure 4.**
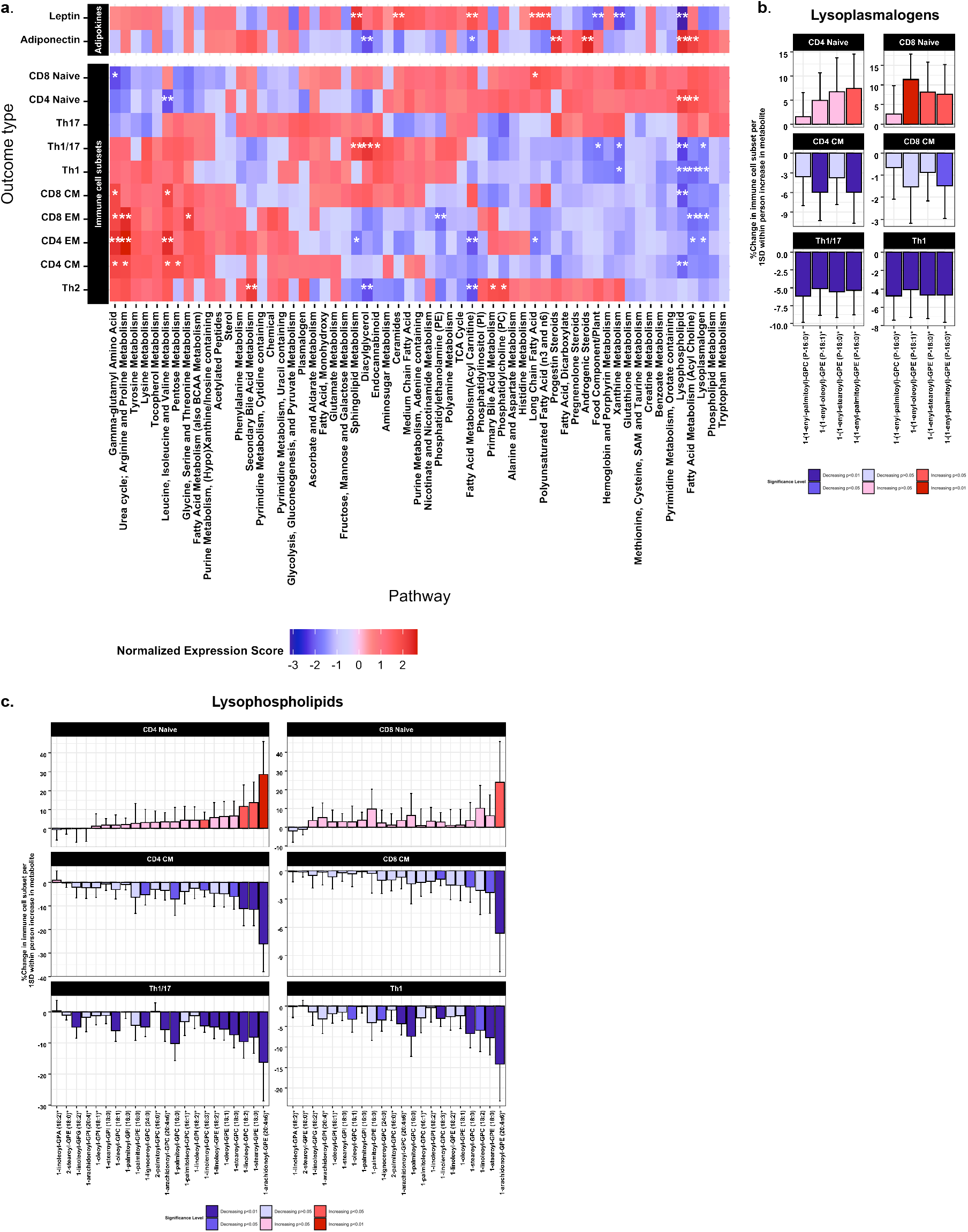
**a**. The heatmap displays the NES from MSEA incorporating results from within person change in metabolite models as they relate to changes in adipokines and immune cell subsets for individuals randomized to intermittent CR. We focused on intermittent CR for these analyses, as our analysis of T cell subsets identified the significant changes for individuals randomized to this type of CR. Darker red colors on the heatmap denote metabolic pathways for which within person increases in metabolites in the pathway were associated with within person increases in a given T cell subset. Darker blue colors on the heatmap denote metabolic pathways for which within person decreases in metabolites in the pathway were associated with within person decreases in a given T cell subset. For example, the dark blue color for the lysophospholipid pathway and CD4+_CM_ suggests that within person change in lysophospholipid pathway metabolites generally are associated with reduction in proportion of CD4+_CM_.*denotes FDR-adjusted pathways with p<0.05 and **denotes FDR-adjusted pathways with p<0.01. **b**. Individual results for metabolites in lysoplasmalogen pathway identified in MSEA as potentially mediating some of the association between intermittent CR and change in selected T cell subsets. Bars are colored based on their direction and level of significance in individual analyses. **c**. Individual results for metabolites in lysophospholipid pathway identified in MSEA as potentially mediating some of the association between intermittent CR and change in selected T cell subsets. Bars are colored based on their direction and level of significance in individual analyses.

## Discussion

This randomized controlled feeding study assessed the effects of intermittent or daily CR diets in people with MS on several relevant classes of potential biologic mediators that included adipokines, various T cell subsets and the circulating metabolome. Leptin and adiponectin did not change significantly in individuals randomized to either CR diet, while randomization to an intermittent CR diet was associated with alterations in T cell subsets and in several classes of biologically relevant lipid metabolites.

Our evaluation of changes in T cell subsets associated with intermittent fasting including notable proportional increases in naïve cells and decreases in memory cells. The observed changes in naïve T cells are consistent with EAE studies of intermittent CR, in which the relative number of naïve T cells increases with fasting in addition to a shift to T regulatory cells (T regs) away from effector T cells (including a reduction in Th1 cells, as in our study).(8) In people, a previous study randomized 17 people with MS recovering from a relapse to alternate-day fasting or usual diets for a period of 15 days.(6) Intermittent CR was associated with decreases in naïve CD4+ and CD8+ T cells (in contrast to our study) and with increased Treg suppressor function (though Treg number was unchanged). However, it is important to note that all participants in that study were treated with glucocorticoids, which could have influenced lymphocyte composition. Additionally, the shorter duration of the relapse recovery study (15 days) may also have played a role in the disparate results compared to our study.

Unexpectedly, we found no change in leptin or adiponectin occurring as the result of randomization to intermittent CR, despite several previous studies implicating a link between fasting and lowering of leptin levels and increasing of adiponectin.(9, 20–22) This finding could be related to our relatively limited sample size and inherent variability of circulating levels (e.g., large variability in levels for participants in the control arm for whom no significant changes in weight were observed. It’s also possible that different fasting protocols (alternate day fasting, time restricted feeding) beyond the 5:2 diet evaluated here may have differential effects on adipokines or other immune cell parameters.(23) Previous mechanistic studies have also demonstrated a link between leptin and T cell activation and pro-inflammatory polarization, which suggested that leptin may mediate the effects of fasting on the relative T cell distribution and function. Our results suggest potentially non-leptin dependent effects of intermittent CR on changes to T cell subsets. As changes in the availability of certain lipid metabolites can shift the distribution of T cells away from effector T cells, it’s possible that some of the observed changes in lipid metabolites may mediate some of the associations noted between intermittent CR and the changes in the relative composition of T cells. An alternative explanation is that reduced levels of glycerophospholids and lysoplasmalogens, may influence functioning of subpopulations of T cells (such as invariant or diverse Natural Killer T cells) that can be activated by these endogenous lipids.(24, 25) Since these T cell populations can then produce other pro-inflammatory mediators that can impact overall T cell function, this offers a potential explanation for the immunological changes observed in our study.

While triglycerides constitute the primary source of stored lipids in humans, phospholipids are critical components of plasma membranes and eicosanoids (e.g., prostaglandins or leukotrienes). Our findings highlight reductions in many classes of glycerophospholipids, including phosphatidylcholine metabolites, among individuals randomized to either CR diet. We also noted reduction in the levels of circulating plasmalogens; plasmalogens are a subtype of glycerophospholipids that may function as endogenous antioxidants and protect membrane lipids or lipoproteins from aberrant reactive oxygen species.(26) However, emerging studies also report other potential functions including involvement in signaling pathways or cell differentiation, which could potentially explain our findings.(26, 27) In particular, we observed a reduction in plasmalogen phosphatidylethanolamines rather than plasmalogen phosphatidylcholines, potentially related to alterations in enzymes necessary for their formation in the peroxisome.

Many of our metabolomic findings are consistent with previous studies of acute CR (over 1 day), prolonged fasting (over 10 days) and following leptin (a known anorectic) replacement therapy in individuals with congenital leptin deficiency.(28–30) For example, one study compared metabolomic profiles in a cohort of individuals over the course of four days: at baseline, following two days in which participants reduced calorie intake by 90%, and a final day in which participants ate ad libitum. Investigators noted significant increases in acyl carnitine metabolites as well as similar changes in glycerophospholipid, lysophospholipids (products of glycerophospholipid metabolism), plasmalogens, and lysoplasmalogens (products of plasmalogen metabolites) as we observed in our study.(29) Similar changes in these lipid classes were also observed in a study measuring metabolomic profiles over the course of a 10-day fast(28) and following leptin replacement therapy among individuals with congenital deficiency.(30) Intriguingly, both studies of acute and prolonged CR also note similar reductions of plasmalogen phosphatidylethanolamines and not phosphatidylcholines associated with fasting.

Strengths of our study include its randomized design and standardization of diet across randomization arms; participants were instructed to consume only study provided foods for a period of 8 weeks, minimizing potential effects related to differences in underlying dietary composition. Furthermore, dietary studies in which food is provided may have better overall adherence (regardless of randomization arm) when compared to guidance-only dietary studies.(31, 32) We also considered a relatively wide array of potential biologic intermediates ranging from immune cells to novel, highly sensitive measures of metabolic health.

This study also has several limitations that are worth noting. First, the study was relatively short in duration; MS is a disease that evolves over decades, so it’s unclear whether what the long-term implications of our findings are. Further, we included relatively few individuals; however, our study was the first of its kind in people with MS and can inform the design of future work in this area. Our feeding study was also remote, and information on adherence was self-reported. In a traditional feeding study, participants consume all meals on-site, and uneaten foods can be more readily monitored by study staff; this was not feasible for the population studied, as most people with relapsing remitting MS are employed and cannot spend their days at a feeding center.(33) As previously noted, adherence to the study diets differed across treatment arms, and, while we attempted to account for this difference analytically, it still possible that residual confounding could have impacted our ability to detect significant changes in biologic mediators.(3)

In summary, we have expanded upon our previous work, demonstrating CR diets are safe and effective ways to achieve weight loss in people with MS, to show that 1) CR is associated with changes in circulating levels of several relevant lipid metabolites, including acyl carnitines and subsets of glycerophospholipids and 2) intermittent CR specifically is associated with a reduction in memory T cell subsets that may be mediated by changes in specific types of glycerophospholipids. Future larger studies are needed to replicate our findings as well as disentangle the underlying mechanisms. This knowledge will improve our understanding of the long-term implications of CR in people with MS, including whether the noted biological changes are relevant to the inflammatory or neurodegenerative aspects of the disease course.

## Methods

### Study Population

As described previously, the ATAC-MS study recruited people with MS aged 18 to 50 from the Johns Hopkins MS Center beginning in December 2015 (clinicaltrials.gov: NCT02647502).(3) Eligible participants met the 2010 criteria for relapsing-remitting MS, had a new lesion or relapse in the past 2 years, had a disease duration ≤ 15 years, Expanded Disability Status Scale (EDSS) <6.0, were stable on an injectable MS therapy or not on any therapy, and were not planning to change vitamin D or thyroid medication for the next 48 weeks. No participants changed their MS therapy over the 8-week period. Participants were also required to have a BMI of at least 23, maintain a stable weight (±8 pounds) for the 3 months preceding the study and not be currently following a specialized diet for MS or other reasons. They also could not smoke >1 cigarette per day. Exclusion criteria were a history of diabetes, eating disorders, kidney disease, warfarin, major surgery in preceding 3 months, chemotherapy in the past year and pregnancy or breast feeding

### Study Design

An overview of the study design is provided in **Figure 1A**. To determine energy needs precisely, all participants underwent indirect calorimetry. Height and weight (used to calculate body mass index [BMI] as kg/m^2^) were also recorded. Randomization was stratified by obesity status (defined as BMI ≥ 30 kg/m^2^). Participants were randomized to 1 of 3 diets: 1) a control diet, in which the participant received 100% of his or her calorie needs 7 days per week, 2) a daily CR diet, in which the participant received 78% of his or her calorie needs 7 days per week, or 3) an intermittent CR diet, in which the participant received 100% of his or her calorie needs on 5 days per week and 25% of his or her calorie needs 2 days per week (i.e., a “5:2” style diet). The composite weekly calorie deficits were similar between the two CR diets. Participants also provided blood samples at baseline and during weeks 4 and 8. Plasma was isolated using a standardized protocol and stored at -80 C, and peripheral blood mononuclear cells (PBMCs) were isolated also following a standardized protocol and stored in liquid nitrogen. Visits during weeks 4 and 8 took place following a 25% intake day for individuals in the intermittent CR arm. All participants provided written consent at enrollment, and the study was approved by the Johns Hopkins Institutional Review Board.

### Intervention

Each of the study diets was standardized to the US median intakes for carbohydrate, protein and fat (to avoid co-intervention due to drastic dietary component changes). The relative macronutrient content over a 1-week period was also comparable between study diets. Over an 8-week period, all meals were prepared by the US Department of Agriculture (USDA) Beltsville Human Nutrition Research Laboratory (Beltsville, MD) and shipped to participants’ homes two times per week. All meals were tailored to the individual participant’s specific caloric needs (as determined by indirect calorimetry and by randomization status). We assessed adherence via in-person 24-hour dietary recalls conducted by study dieticians twice during weeks 4 and 8. We created a continuous measure of adherence as the difference between study-provided calorie intakes and participant-reported intake collected during the 24-hour recalls.

### Outcomes

The primary outcomes of the study were safety and feasibility of different types of CR diets in people with MS; results for the primary outcome were previously reported. For this study we considered the following sets of intermediate outcomes: 1) adipokines, 2) immune cell subsets, and 3) untargeted metabolomics.

#### Adipokines

Plasma leptin and adiponectin were measured using commercially available ELISA kits and were conducted in a single batch.

#### Immune cell subsets

PBMCs from baseline and week 8 were isolated by Ficoll separation and cryopreserved. Cell were thawed and washed in c-RPMI medium. They were stained with antibodies for the following antigens – CD3, CD4, CD8, CD45RO, CCR7, CCR4, CCR6 and CXCR3 and acquired on a MACS Quant flow-cytometer (Miltenyi, Biotec), as previously described.(34, 35) An overview of the gating strategy is provided in **Supplemental Figure 2a**. We identified CD4^+^ and CD8^+^ T cells quantified as percent CD3, which reflected percent of viable cells. Within these we identified memory subsets including effector memory (T_EM_: CCR7-CD45RO+), central memory (T_CM_: CCR7+ CD45RO+), and naïve cells (T_naive_: CCR7+ CD45RO-). As defined by CCR4, CCR6, and CXCR3, we also quantified (as %CD4+): Th2 (CCR4+, CCR6-, CXCR3-), Th1 (CCR4-, CCR6+, CXCR3-), Th17 (CCR4-, CCR6+, CXCR3-) and Th1/17 (CCR4-, CCR6+, CXCR3+). **Supplemental Figure 2b** also includes s select fluorescence minus one (FMO) staining controls.

#### Metabolomics

Blood samples provided at (at baseline and weeks 4, 8) were used to obtain plasma. For individuals randomized to intermittent CR, samples at weeks 4 and 8 were collected following a 25% calorie-needs day (i.e., following a fasting day). Untargeted metabolomics was performed in a single batch at Metabolon (Durham, NC), and methods have been described in detail elsewhere.(18, 19) Briefly, samples were thawed and underwent additional preparation (derivatization). The derivatized samples were subjected to either gas chromatography followed by mass spectrometry (GC/MS) or liquid chromatography followed by tandem mass spectrometry (LC/MS/MS). Mass spectra obtained from these techniques were then matched to a library of spectra derived from standards to identify specific metabolites, and the area under the curve for the mass spectra was used to calculate the relative abundance of each metabolite. We initially included 795 metabolites. We then implemented the following quality control (QC) procedure to identify and remove potential outlying metabolites. First, we removed metabolites with >20% missing values across samples (n=125) and imputed missing metabolite values using k-nearest neighbors (10 neighbors used for each imputation) for the remaining 670 metabolites; consistent results were observed when using the minimum value of observed metabolites. We then log-transformed and scaled all metabolites. Our previous results demonstrate excellent reproducibility for metabolomics in that we have shown intraclass correlation coefficients (ICC) to be high (median ICCs across metabolites for replicate samples was 0.94; 95% CI: 0.86, 0.98 and 1.2% of metabolites had ICCs <0.40).(18)

### Statistical analysis

We evaluated differences in the overall change in each of the intermediate outcomes (immune cell subtype, metabolic trait, or metabolite) associated with randomization to CR diets, according to the intention to treat (ITT) principle and using mixed effects regression models. We also considered models comparing randomization to any CR diet (daily or intermittent) with control diets. Models were additionally adjusted for age, sex, MS disease-modifying therapy, and adherence to study diets. We fit a separate model for each immune cell type or metabolic trait. For metabolomics analyses, since our sample size was relatively small, we performed analyses concentrating on differences in overall metabolic pathways (to reduce the number of statistical tests) as our primary analyses for this outcome. To do so, we classified metabolites into groups (>3 metabolites) based on related biologic function (e.g., carnitine metabolism, tryptophan metabolism, among others); pathway memberships for each metabolite are included as a part of **Supplemental Table 1**. We then conducted a metabolite-set enrichment analysis (MSEA). Like its predecessor gene set enrichment analysis (GSEA), MSEA is a computational method that evaluates whether sets of metabolites (or genes, in the case of GSEA) demonstrates concordant differences in a comparison of different biological states.(36, 37) For each metabolic pathway, a normalized enrichment score (NES) is calculated using similar methods to GSEA in that that each pathway NES is normalized by the size of the pathway and incorporates individual statistical tests for each of the metabolites considered. Here, we considered the rate of change for each metabolite estimated from mixed effects models divided by its standard error as the tests supplied to MSEA. Statistical significance for each pathway is then assigned using a permutation procedure to generate the null distribution of the NES, and the p value is calculated relative to this null distribution as in GSEA.(36) In secondary metabolomics analyses, we also considered the results of changes in individual metabolites.

In exploratory analyses, we also assessed whether changes in adipokines or immune cell subtypes were mediated by within-person changes in circulating metabolite levels. To do so, we calculated an overall person-specific metabolite mean (e.g., the average of a person’s metabolite levels across all visits) and a within-person measure as the difference between the overall person-specific mean of a metabolite and the person’s metabolite level at each time point. We then fit a mixed model for a given intermediate outcome (adipokine or T cell subset) in which we included a term for overall metabolite mean and a term for the within-person difference as well as similar set of covariates as in our primary analyses (i.e., age, sex, MS DMT, and adherence). Then, similar to overall metabolomics-based analyses, we performed MSEA on the estimated rate of within-person change for each metabolite divided by its standard error. We also fit within-person change models for changes in adipokines as they relate to T cell subsets; changes in adipokines have been linked with differences in T cell function in previous studies.(6, 7, 38) We adjusted all analyses for multiple comparisons using the false discovery rate (FDR).

## Data Availability

All data produced in the present study may be available upon reasonable request to the authors with appropriate data sharing requests in place.

---

**Supplemental Figure 1.**
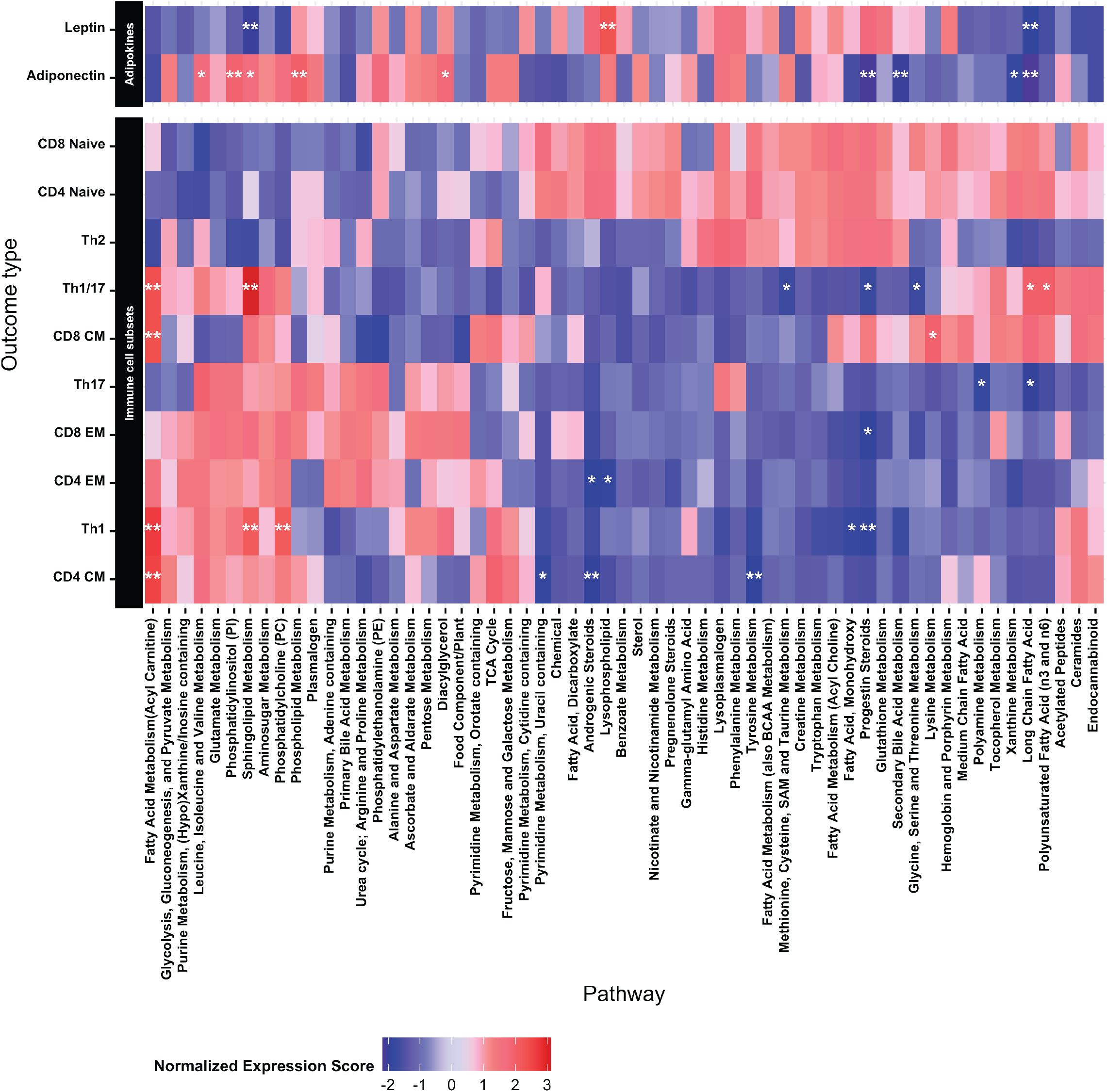
The heatmap displays the NES from MSEA incorporating results from within person change in metabolite models for individuals randomized to daily CR as they relate to changes in adipokines and immune cell subsets. Darker red colors on the heatmap denote metabolic pathways for which within person increases in metabolites in the pathway were associated with within person increases in a given T cell subset. Darker blue colors on the heatmap denote metabolic pathways for which within person decreases in metabolites in the pathway were associated with within person decreases in a given T cell subset. *denotes FDR-adjusted pathways with p<0.05 and **denotes FDR-adjusted pathways with p<0.01.

**Supplemental Figure 2.**
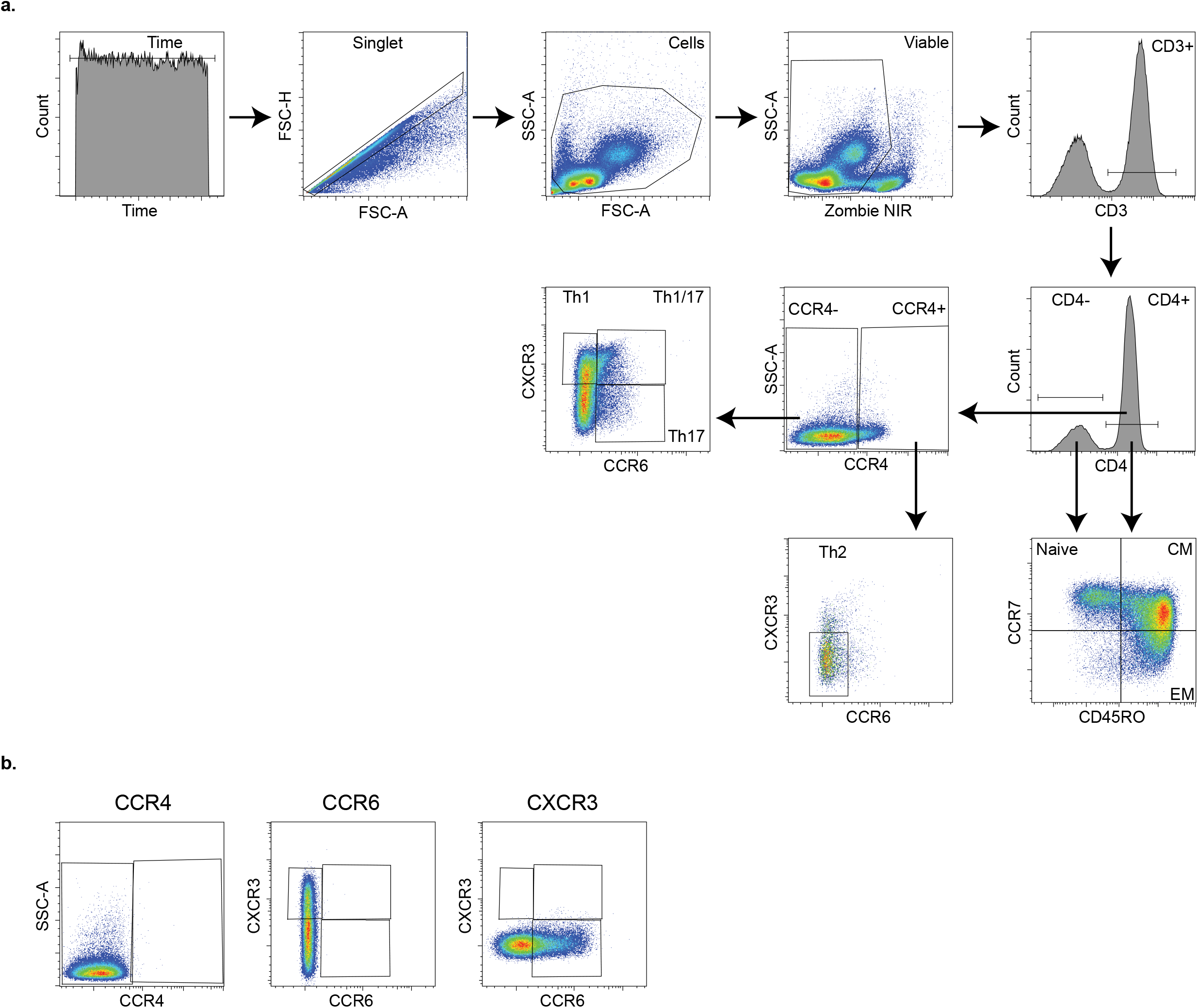
**a**. Gating strategy used to identify T cell subsets. **b**. Select fluorescence minus one (FMO) staining controls for immunophenotyping experiments.

## Conflicts of interest

Dr. Fitzgerald, Dr. Bhargava, Mr. Smith, Ms. Henry-Barron, Ms. Vizthum, Dr. Cassard, Dr. Kappogiannis, Mr. Sullivan, and Dr. Baer report no disclosures

Dr. Kornberg has received consulting fees from Biogen Idec, Janssen Pharmaceuticals, Novartis, OptumRx, and TG Therapeutics.

Dr. Calabresi has received consulting fees from Disarm, NervGen, and Biogen and is PI on grants to JHU from Genentech.

Dr. Mowry has grants from Biogen, is site PI for studies sponsored by Biogen and Genentech, has received free medication for a clinical trial from Teva, and receives royalties for editorial duties from UpToDate.

## Author contributions

**Designing research studies:** KCF, PB, PAC, EMM

**Conducting experiments:** PB, MDS, DK

**Acquiring data:** KCF, BHB, DV, SDC, PS, DB, EMM

**Analyzing data:** KCF, MDS, PB, MDK

**Manuscript preparation and editing:** All authors

## Acknowledgements

The authors would like to acknowledge advice and guidance provided the late Dr. John Milner of the United States Department of Agriculture in developing the design of the study. This publication was made possible by the Johns Hopkins Institute for Clinical and Translational Research (ICTR) which is funded in part by Grant Number UL1 TR 001079 from the National Center for Advancing Translational Sciences (NCATS) a component of the National Institutes of Health (NIH), and NIH Roadmap for Medical Research. Its contents are solely the responsibility of the authors and do not necessarily represent the official view of the Johns Hopkins ICTR, NCATS or NIH. This study was supported in part by the Intramural Research Program of the National Institute on Aging and support from a Catalyst Award from Johns Hopkins University.

## Notes

### Clinical Trial

NCT02647502

### Author Declarations

A Johns Hopkins Institutional Review Board

